# The impact of lockdown measures on COVID-19: a worldwide comparison

**DOI:** 10.1101/2020.05.22.20106476

**Authors:** DI Papadopoulos, I Donkov, K Charitopoulos, S Bishara

**Author notes:** Corresponding author Samuel Bishara, Division of surgery, West Middlesex University Hospital, Chelsea and Westminster NHS Trust, Twickenham Road, London TW7 6AF, Email.

## Abstract

**Objective:** We aimed to determine which aspects of the COVID-19 national response are independent predictors of COVID-19 mortality and case numbers.

**Design:** Comparative observational study between nations using publicly available data.

**Setting:** Worldwide Participants Covid-19 patients

**Interventions:** Stringency of 11 lockdown policies recorded by the Blavatnik School of Government database and earliness of each policy relative to first recorded national cases

**Main outcome measures:** Association with log_10_ National deaths (LogD) and log_10_ National cases (LogC) on the 29^th^ April 2020 corrected for predictive demographic variables

**Results:** Early introduction was associated with reduced mortality (n=137) and case numbers (n=150) for every policy aside from testing policy, contact tracing and workplace closure. Maximum policy stringency was only found to be associated with reduced mortality (p=0·003) or case numbers (p=0·010) for international travel restrictions. A multivariate model, generated using demographic parameters (r^2^=0·72 for LogD and r^2^=0·74 for LogC), was used to assess the timing of each policy. Early introduction of first measure (significance p=0·048, regression coefficient β=-0·004, 95% confidence interval 0 to -0·008), early international travel restrictions (p=0·042, β=-0·005, -0·001 to - 0·009) and early public information (p=0·021, β=-0·005, -0·001 to -0·009) were associated with reduced LogC. Early introduction of first measure (p=0·003, β=-0·007, -0·003 to -0·011), early international travel restrictions (p=0·003, β=-0·008, -0·004 to-0·012), early public information (p=0·003, β=-0·007, 0·003 to -0·011), early generalised workplace closure (p=0·031, β=-0·012, -0·002 to -0·022) and early generalised school closure (p=0·050, β=-0·012, 0 to -0·024) were associated with reduced LogC.

**Conclusions:** At this stage in the pandemic, early institution of public information, international travel restrictions, and workplace closure are associated with reduced COVID-19 mortality and maintaining these policies may help control the pandemic.

**What is already known on this topic:** The COVID-19 pandemic has spread rapidly throughout the world and presented vast healthcare, economic and political challenges. Many nations have recently passed the peak of their infection rate, and are weighing up relaxation of lockdown strategies. Though the effect of individual lockdown policies can be estimated by modelling, little is known about the impact of individual policies on population case numbers or mortality through comparison of differing strategies between nations. A PubMed search was carried out on the 14/5/20 using keywords including “novel coronavirus-infected pneumonia”, “2019-nCoV”, “Sars-Cov-2”, “Covid-19”, “lockdown”,” policy”, “social distancing”, “isolation”, “quarantine” and “contact tracing” returned 258 studies in total. Following scanning of the above results, we found 19 studies that have examined the effect of lockdown within a region, which have demonstrated a reduction in case numbers after the introduction of a lockdown. There are no previous studies that have compared the effectiveness of government lockdowns between nations to determine the effectiveness of specific policies.

**What this study adds:** This study examines the corollary between government policy and COVID-19 case numbers and mortality, correct as of the 29th of April 2020, for every nation that there is available date within the Blavatnik School of Government database on COVID-19 policy. The study demonstrates that early generalised school closure, early generalised workplace closure, early restriction of international travel and early public information campaigns are independently associated with reduced national COVID-19 mortality. The maximum stringency of individual lockdown policies were not associated with reduced case numbers or mortality. Early reintroduction of these policies may be most effective in a relapse of the pandemic, though, school closure, workplace closure and restriction of international travel carry heavy politico-economic implications. There was no measurable effect of maximum stringency of lockdown policy on outcome at this point in time, indicating that early timing of lockdown introduction is of greater importance than its stringency, provided that the resultant viral reproductive rate is less than 1.

## Introduction

The Sars-CoV-2 outbreak started initially in Wuhan where the first case of novel coronavirus was reported in December 2019. It has spread rapidly to almost 185 countries and caused thousands of deaths. Currently the number of confirmed cases is increasing worldwide and the pressure on global healthcare systems has been considerable, with more than 3 million cases and 220 thousand deaths by the end of April 2020.^1^ As the current testing methods are poor, the true incidence of the disease is estimated to be much higher.^1,2^

Person to person spread may be direct, where virus particles emitted through respiratory droplets on coughing, sneezing or talking enter the respiratory tract of those nearby^3^, or indirect, from contact with contaminated surfaces. The viral viability on surfaces has been studied and tested positive for up to 24 hours on cardboard and 72 hours on plastic and steel.^4^

Symptoms of fever, cough, dyspnoea, fatigue, headache, diarrhoea^5^ typically appear after a short incubation period of 5·2 days^6^ and the mean period to fatal events is 14 days from the symptoms’ initiation. A reasonably high rate of asymptomatic carriage is noted with 20% of the cases being completely asymptomatic and 30% afebrile.^7^ Transmission of infection can occur before the onset of symptoms.^8^ These two factors have made it more difficult to track the pandemic.

Whilst the interconnectedness of the modern world has been a catalyst for the unprecedented rapidity with which COVID-19 has spread around the globe, there has never before been the means to monitor and control a pandemic that we have available today. Significant efforts have been made towards the development of vaccines or therapeutic protocols.^8^ However, In the absence of highly effective treatments, a contagious novel disease still has enormous potential for morbidity, and social distancing, allied with contact tracing, remains the principle control strategy,^9,10^ albeit one that comes with considerable economic and political challenges.^11^

Governmental responses exhibit significant nuance and heterogeneity based on their own projections^12^ and have been constructed according to differences in case numbers, deaths ratios, local health system strengths and limitations along with financial support.^13^

Social distancing policies include school and workplace closure, cancellation of public events and gatherings in addition to movement restrictions with public transport closure, and prohibition of internal and international travel. The University of Oxford, Blavatnik School of Government has contributed to our understanding by meticulously documenting these policies according to date, based on news articles or government press releases, and categorizing them into specific groups, each graded according to policy stringency and their application as targeted or general population strategies.^13^ Likewise, global data for active cases, incidence rate and case fatality ratio has been recorded by John Hopkins University. This database was combined with the Blavatnik School of government database together with demographic factors in the studied countries.^1,13,14^

The present study aimed to incorporate every country in the world, data permitting, to examine the impact of differing lockdown approaches with regards to their strength and timing to determine their correlation with case numbers and the mortality rates. This was assessed with demographic variables which could be predictive of the COVID-19 outbreak. Though there are pitfalls in comparing outcomes between countries, we believe this methodology is relevant, as this study indicates that the pandemic mortality can be accurately modelled with a small number of pre-pandemic demographic characteristic. When these are factored, the response to the pandemic can be brought into focus. Whilst there are a number of studies that have evaluated the effect of lock down in specific region,^11,15–17^ we are not aware of any studies that have evaluated the effect of individual government social distancing policy on the pandemic before by making a comparison across all nations (data permitting). At this time many countries are evaluating relaxation of their policies and this study may provide useful information on government policy, indicating the most effective lockdown policies to control virus spread,^18^ and their timing, should they need to be reintroduced in the event of a relapse.^19^

## Methods

### Data collection

COVID-19 morbidity data was obtained from the John Hopkins university public database.^1^ Data for mortality, case numbers, test numbers and date of first case and death were obtained for all countries that had COVID-19 cases on the 29^th^ April 2020. The primary measures of national epidemic used were log base 10 of total cases (LogC) and log base 10 of total deaths (LogD), correct as of April 29th 2020. Log base 10 was used as the resultant parameters had a more normal distribution. Dates were counted in days over a period of 120 days from 1st of January, day 1, to 29th of April, day 120. The date of first death was fixed at 121 for the 13 countries that did not have a death during the analysis period.

Publically available data on the national response to the COVID-19 pandemic was extracted from the Oxford COVID-19 Government Response Tracker, Blavatnik School of Government database.^13^ Data is present for 151 nations for 11 policies; C1 school closure, C2 workplace closure, C3 cancelling of public events, C4 restriction on gatherings, C5 closure of public transport, C6 stay at home restrictions, C7 domestic travel restrictions, C8 international travel restrictions, H1 public information, H2 testing framework and H3 contact tracing. Policies C3, C5, H1, and H3 are scored on an ordinal scale from 0-2. Policies C1, C2, C6, C7 and H2 are scored 0 to 3. Policies C4 and C8 are scored 0-4. As an example, the school closure index C1 was categorized as 0: no measures, 1: recommend closing, 2: Require closing (only partially) 3: Require closing (all levels). Additionally, policies C1-C7 and H1 were categorised on a binary scale if they were applied as generalised policies for the entire populace -1 or selective policies - 0. Where data was absent this was scored as 0. A composite stringency index, generated by averaging the percentage of maximum score for 9 policies; C1-C8 and H1 on each day was also evaluated. This index has a range from 0-100. Data was extracted for every day from the 1st of January 2020 to April 29th 2020, for 151 nations. One country had no COVID-19 cases and was excluded from the analysis. The policy introduction date was likewise counted in days from the 1st of January for the same follow up period of 120 days. The latest a COVID-19 first case was recorded for any country was on day 92. Therefore, the shortest follow up period was 28 days. In order to include all countries in analysis, whether they had introduced one of the lock down policies or not, the relative earliness of lockdown introduction was defined as the earliest date of lockdown policy counted back form 28 days after the first recorded case within a country (e.g. If no lockdown policy was introduced, or introduced > 28 days after the first case, this was recorded as 0. If a policy was introduced 28 days after the first case this was recorded as 1. If introduced on the day of the first case this was recorded as 29 etc). This value was assessed against LogC and LogD by bivariate correlation and multivariate linear regression.

Demographic data for country area, population, density, average purchasing power, number of flights per year was obtained from the World Bank; average household size was obtained from the United Nations database.^14^

### Analysis

Data was collated in a single database. All statistical analysis was carried out by IBM SPSS. The maximum stringency index, the individual policy maximum stringency on an ordinal scale and the maximum binary score for generalisation of policy, were compared with LogC or LogD, by bivariate correlation, assessed by Rho Spearman’s coefficient. The relative timing of policy introduction, as described above, was assessed against LogC and LogD using Pearson’s correlation.

A multivariate regression model was created for assessment of the independent determination of LogC and LogD by policy timing, for every country (data permitting). The model was assessed by ANOVA and variable coefficients were assessed for significance (P ≤0·05) by t test. Firstly, predictive demographic factors for LogC and LogD were determined. The following factors were assessed; flight arrival numbers, population density, log population, continent, area, average household size, purchasing power, date of first case, date of first death, longitude, latitude, life expectancy. The factors were reduced by iteration with backwards stepwise regression, including factors with P <0·05 and eliminating factors with p >0·1. The final demographic model was assessed with the timing of introduction of each lockdown measure in turn, in separate multivariate regressions, for prediction of LogC and LogD.

### Patient and public Involvement statement

Patients and the public were not involved in the research.

## Results

Table 1 demonstrates a summary of the demographic data used in the multivariate regression model and the outcome measures; LogC and LogD. 150 countries were included in the database. LogD was recorded for the 137 countries that had 1 or more death as of 29^th^ of April 2020. Smoking rates and flight number were also associated with LogC and LogD, but not used in the subsequent multivariate model due to missing data. Longitude was strongly associated with outcome on multivariate regression but not on bivariate association here. Earlier first case and first death were strongly correlated with LogC (n=150) and LogD (n=137).

**Table 1:** Univariate correlation with log_10_ Total cases and log_10_ Total Deaths for evaluated demographic factors. *= P≤0.05, **= P ≤0.01.

|  |  |  | Log Total Cases |  |  | Log Total Deaths |  |  |
| --- | --- | --- | --- | --- | --- | --- | --- | --- |
|  | Mean | 95% CI | Pearson Correlation (95% CI) | Sig. (2-tailed) | N | Pearson Correlation (95% CI) | Sig. (2-tailed) | N |
| Log Population | 6.99 | 0.14 | 0.509**<br>(0.394, 0.609) | 0.000 | 150 | 0.532**<br>(0.422, 0.626) | 0.000 | 137 |
| Density | 397.06 | 297.06 | -0.070<br>(-0.193, 0.126) | 0.394 | 150 | -0.112<br>(-0.191, 0.05) | 0.193 | 137 |
| Area km2 | 864100 | 343485 | 0.326**<br>(0.174, 0.451) | 0.000 | 150 | 0.336**<br>(0.189, 0.490) | 0.000 | 137 |
| Longitude | 18.56 | 9.55 | -0.012<br>(-0.189, 0.162) | 0.882 | 150 | -0.070<br>(-0.236, 0.093) | 0.419 | 137 |
| Household size | 3.82 | 0.22 | -0.445**<br>(-0.569, -0.307) | 0.000 | 134 | -0.442**<br>(-0.554, -0.315) | 0.000 | 123 |
| Flight numbers | 8522548 | 2420254 | 0.626**<br>(0.534, 0.704) | 0.000 | 136 | 0.666**<br>(0.548, 0.754) | 0.000 | 125 |
| Smoking percentage | 20.17 | 1.59 | 0.328**<br>(0.172, 0.461) | 0.000 | 139 | 0.343**<br>(0.220, 0.466) | 0.000 | 128 |
| Life expectancy | 74.53 | 1.16 | 0.527**<br>(0.414, 0.626) | 0.000 | 150 | 0.465**<br>(0.347, 0.566) | 0.000 | 137 |
| Purchasing power | 1.58 | 0.36 | 0.242**<br>(0.126, 0.484) | 0.003 | 150 | 0.187*<br>(0.093, 0.418) | 0.029 | 137 |
| 1<sup>st</sup> death | 83.68 | 2.96 | -0.560**<br>(-0.717, -0.470) | 0.000 | 150 | -0.600**<br>(-0.713, -0.469) | 0.000 | 137 |
| 1<sup>st</sup> Case | 62.68 | 3.23 | -0.649**<br>(-0.743, -0.515) | 0.000 | 150 | -0.605**<br>(-0.719, -0.469) | 0.000 | 137 |
| Log Total Cases | 3.12 | 0.18 | - |  |  | 0.920<br>(0.886, 0.944) | 0.000 | 137 |
| Log Total Deaths | 1.75 | 0.19 | 0.920<br>(0.886, 0.944) | 0.000 | 137 | - |  |  |

Table 2 shows bivariate correlation of lockdown policy versus LogC or LogD. The Rho Spearman Coefficient was used throughout. There was no consistent association between the maximum strength of the overall index, maximum stringency of any policy, or generalisation of any policy and reduction in LogC or LogD. In fact, the opposite was seen in some cases; generalisation of C2, generalisation of C7, and increased stringency of H2 and H3 were associated with increased LogC. Likewise, generalisation of C6 and increased stringency and generalisation of C7 were associated with increased LogD. Maximum stringency of policy was only found to be associated with reduced LogC (p=0·010) and LogD (p=0·003) for one policy: international travel restrictions.

**Table 2:** The correlation between maximum stringency of policy versus Log_10_ Total cases and Log_10_ Total Deaths. ^*^= P≤0.05, ^**^= P ≤0.01.

| Policy (Ordinal scale range) |  |  | Log Total Cases |  | Log Total Deaths |  |
| --- | --- | --- | --- | --- | --- | --- |
|  | Mean | 95% CI | Spearman Correlation (95% CI) | Sig. (2-tailed), n=150 | Spearman Correlation (95% CI) | Sig. (2-tailed) n=137 |
| **C1 school closing (0-3)** | 2.92 | 0.07 | 0.048<br>(-0.158, 0.236) | 0.559 | 0.057<br>(-0.167, 0.256) | 0.511 |
| **C1 general policy (0-1)** | 0.97 | 0.03 | -0.03<br>(-0.256, 0.218) | 0.719 | -0.025<br>(-0.246, 0.211) | 0.776 |
| **C2 work place closing (0-3)** | 2.48 | 0.15 | 0.086<br>(-0.062, 0.240) | 0.295 | 0.082<br>(-0.112, 0.221) | 0.342 |
| **C2 general policy (0-1)** | 0.84 | 0.06 | 0.139<br>(0.014, 0.298) | 0.091 | 0.143<br>(-0.009, 0.301) | 0.096 |
| **C3 cancel public events (0-2)** | 1.94 | 0.05 | 0.149<br>(-0.081, 0.306) | 0.070 | 0.165<br>(-0.039, 0.312) | 0.054 |
| **C3 general policy (0-1)** | 0.94 | 0.04 | 0.037<br>(-0.161, 0.249) | 0.656 | 0.047<br>(-0.172, 0.251) | 0.589 |
| **C4 restriction on gatherings (0-4)** | 3.22 | 0.20 | 0.102<br>(-0.062, 0.281) | 0.212 | 0.049<br>(-0.120, 0.230) | 0.569 |
| **C4 general policy (0-1)** | 0.83 | 0.06 | -0.037<br>(-0.182, 0.140) | 0.646 | -0.025<br>(-0.188, 0.160) | 0.776 |
| **C5 close public transport (0-2)** | 1.39 | 0.12 | -0.042<br>(-0.215, 0.129) | 0.610 | -0.006<br>(-0.165, 0.167) | 0.948 |
| **C5 general policy (0-1)** | 0.69 | 0.07 | -0.069<br>(-0.232, 0.119) | 0.402 | -0.052<br>(-0.225, 0.120) | 0.545 |
| **C6 stay at home restriction (0-3)** | 1.83 | 0.14 | 0.056<br>(-0.107, 0.225) | 0.497 | 0.034<br>(-0.137, 0.225) | 0.694 |
| **C6 general policy (0-1)** | 0.75 | 0.07 | 0.178*<br>(0.015, 0.339) | 0.029 | 0.197*<br>(0.020, 0.372) | 0.021 |
| **C7 domestic travel restriction (0-3)** | 1.79 | 0.09 | 0.117<br>(-0.055, 0.271) | 0.155 | 0.180*<br>(0.025, 0.344) | 0.035 |
| **C7 general policy (0-1)** | 0.81 | 0.06 | 0.165*<br>(0.006, 0.308) | 0.044 | 0.215*<br>(0.045, 0.371) | 0.012 |
| **C8 international travel restriction (0-4)** | 3.60 | 0.12 | -0.210**<br>(-0.387, -0.013) | 0.010 | -0.249**<br>(-0.419, -0.097) | 0.003 |
| **H1 public information (0-2)** | 1.97 | 0.03 | 0.014<br>(-0.173, 0.189) | 0.862 | 0.035<br>(-0.176, 0.230) | 0.689 |
| **H1 general policy (0-1)** | 1.00 | 0.00 |  |  |  |  |
| **H2 testing policy (0-3)** | 1.62 | 0.13 | 0.193*<br>(0.050, 0.355) | 0.018 | 0.128<br>(-0.062, 0.295) | 0.135 |
| **H3 contact tracing (0-2)** | 1.30 | 0.12 | 0.167*<br>(0.006, 0.316) | 0.041 | 0.090<br>(-0.096, 0.276) | 0.296 |
| **Index maximum (0-100)** | 85.11 | 2.07 | -0.080<br>(-0.250, 0.091) | 0.332 | -0.029<br>(-0.190, 0.136) | 0.735 |

However, for early timing of policy introduction, correlations with reduced LogC and LogD were almost universally noted (Table 3); early timing of first lockdown measure (p=0·000), early school closure C1 (p=0·000) and generalisation of C1 (p=0·000), early generalisation of workplace closure C2 (p=0·000), early cancellation of public events C3 (p=0·000) and early generalisation of C3 (p=0·000), early restriction on gatherings C4 (p=0·000) and early generalisation of C4 (p=0·000), early closure of public transport C5 (p=0·006) and early generalisation of C5 (p=0·000), early stay at home restrictions C6 (p=0·000) and generalisation of C6 (p=0·000), early restriction of domestic movement C7 (p=0·000) and early generalisation of C7 (p=0·000), early international travel restrictions C8 (p=0·000), early public information campaign H1 (p=0·000) and early generalisation of H1 (p=0·000), were found to be associated with reduction in the LogC. When examining Log of the total deaths; early timing of first lockdown measure (p=0·000), early school closure C1 (p=0·000) and generalisation of C1 (p=0·000), early generalisation of work place closure C2 (p=0·002), early cancellation of public events C3 (p=0·000), early generalisation of C3 (p=0·000), early restriction on gathering C4 (p=0·000), early generalisation of C4 (p=0·000), early closure of public transport C5 (p=0·023), early generalisation of C5 (p=0·007), early stay at home restrictions C6 (p=0·000) and generalisation of C6 (p=0·001), early restriction of domestic movement C7 (p=0·000), and generalisation of C7 (p=0·000), early international travel restrictions C8 (p=0·000), early public information H1 (p=0·000) and early generalisation of H1 (p=0·000), were found to be associated with reduction in LogD.

**Table 3:** The correlation between timing of policy introduction versus Log_10_ Total Cases and Log_10_ Total Deaths. The mean value indicates the timing of policy introduction as a countback from 28 days after the first case was introduced. Eg the earliest policy introduced was Public information at (42·1-29) = 13·1 days before first recorded national case and the latest; stay at home restrictions at 19·8 days after the first case. *= P≤0.05, **= P ≤0.01.

|  | mean | 95% CI | Log Total Cases |  | Log Total Deaths |  |
| --- | --- | --- | --- | --- | --- | --- |
|  |  |  | Pearson Correlation<br>95% CI | Sig. (2-tailed),<br>n=150 | Pearson Correlation<br>95% CI | Sig. (2-tailed)<br>n=137 |
| **C1 school closing timing** | 21.1 | 2.1 | -0.599**<br>(-0.705, -0.488) | 0.000 | -0.565**<br>(-0.674, -0.396) | 0.000 |
| **C1 timing of general policy** | 19.9 | 2.1 | -0.584**<br>(-0.711, -0.459) | 0.000 | -0.585**<br>(-0.702, -0.430) | 0.000 |
| **C2 work place closing timing** | 22.7 | 1.7 | -0.157<br>(-0.328, 0.020) | 0.055 | -0.087<br>(-0.250, 0.067) | 0.313 |
| **C2 general policy timing** | 12.3 | 2.2 | -0.330**<br>(-0.465, -0.186) | 0.000 | -0.268**<br>(-0.414, -0.094) | 0.002 |
| **C3 cancel public events timing** | 21.9 | 2.2 | -0.528**<br>(-0.649, -0.392) | 0.000 | -0.474**<br>(-0.609, -0.297) | 0.000 |
| **C3 timing of general policy** | 19.2 | 2.3 | -0.526**<br>(-0.639, -0.390) | 0.000 | -0.467**<br>(-0.596, -0.295) | 0.000 |
| **C4 restriction on gatherings timing** | 12.2 | 2.2 | -0.412**<br>(-0.527, -0.266) | 0.000 | -0.386**<br>(-0.507, -0.242) | 0.000 |
| **C4 timing of general policy** | 11.4 | 2.1 | -0.424**<br>(-0.545, -0.279) | 0.000 | -0.380**<br>(-0.493, -0.238) | 0.000 |
| **C5 close public transport timing** | 12.0 | 2.3 | -0.225**<br>(-0.449, 0.014) | 0.006 | -0.194*<br>(-0.430, 0.035) | 0.023 |
| **C5 timing of general policy** | 9.9 | 2.3 | -0.342**<br>(-0.483, -0.168) | 0.000 | -0.230**<br>(-0.393, -0.058) | 0.007 |
| **C6 stay at home restriction timing** | 9.2 | 1.9 | -0.360**<br>(-0.487, -0.170) | 0.000 | -0.308**<br>(-0.453, -0.118) | 0.000 |
| **C6 general policy timing** | 7.2 | 1.8 | -0.365**<br>(-0.486, -0.210) | 0.000 | -0.292**<br>(-0.434, -0.129) | 0.001 |
| **C7 domestic travel restriction timing** | 14.8 | 1.9 | -0.366**<br>(-0.505, -0.203) | 0.000 | -0.380**<br>(-0.521, -0.204) | 0.000 |
| **C7 general policy timing** | 10.2 | 1.9 | -0.342**<br>(-0.459, -0.195) | 0.000 | -0.321**<br>(-0.464, -0.159) | 0.000 |
| **C8 international travel restriction timing** | 37.1 | 3.9 | -0.454**<br>(-0.579, -0.329) | 0.000 | -0.447**<br>(-0.571, -0.294) | 0.000 |
| **H1 public information timing** | 42.1 | 4.0 | -0.347**<br>(-0.481, -0.197) | 0.000 | -0.397**<br>(-0.538, -0.236) | 0.000 |
| **H1 timing of general policy** | 39.1 | 4.1 | -0.333**<br>(-0.485, -0.173) | 0.000 | -0.375**<br>(-0.506, -0.218) | 0.000 |
| **H2 testing policy timing** | 33.1 | 3.4 | -0.043<br>(-0.203, 0.125) | 0.602 | -0.079<br>(-0.238, 0.116) | 0.359 |
| **H3 contact tracing timing** | 24.0 | 3.1 | -0.019<br>(-0.228, 0.153) | 0.820 | -0.012<br>(-0.179, 0.144) | 0.893 |
| **Index timing** | 48.4 | 3.9 | -0.381**<br>(-0.503, -0.244) | 0.000 | -0.400**<br>(-0.527, -0.236) | 0.000 |

As there was no clear relationship between the maximum strength of policy and reduced mortality, this was excluded from subsequent multivariate analysis, which focused on timing which demonstrated a universal trend for benefit on univariate correlation and no adverse correlations, as all the Pearson coefficients in Table 3 are negative. This is illustrated in Figure 1, where relative timing of school closure is plotted against LogD for 137 countries. A positive value on the x axis is indicative of earlier closure. This shows that a later date is associated with and increased LogD; however when the maximum strength of policy is assessed on a scale from 0-2, increased stringency is associated with a worse outcome.

**Figure 1:**
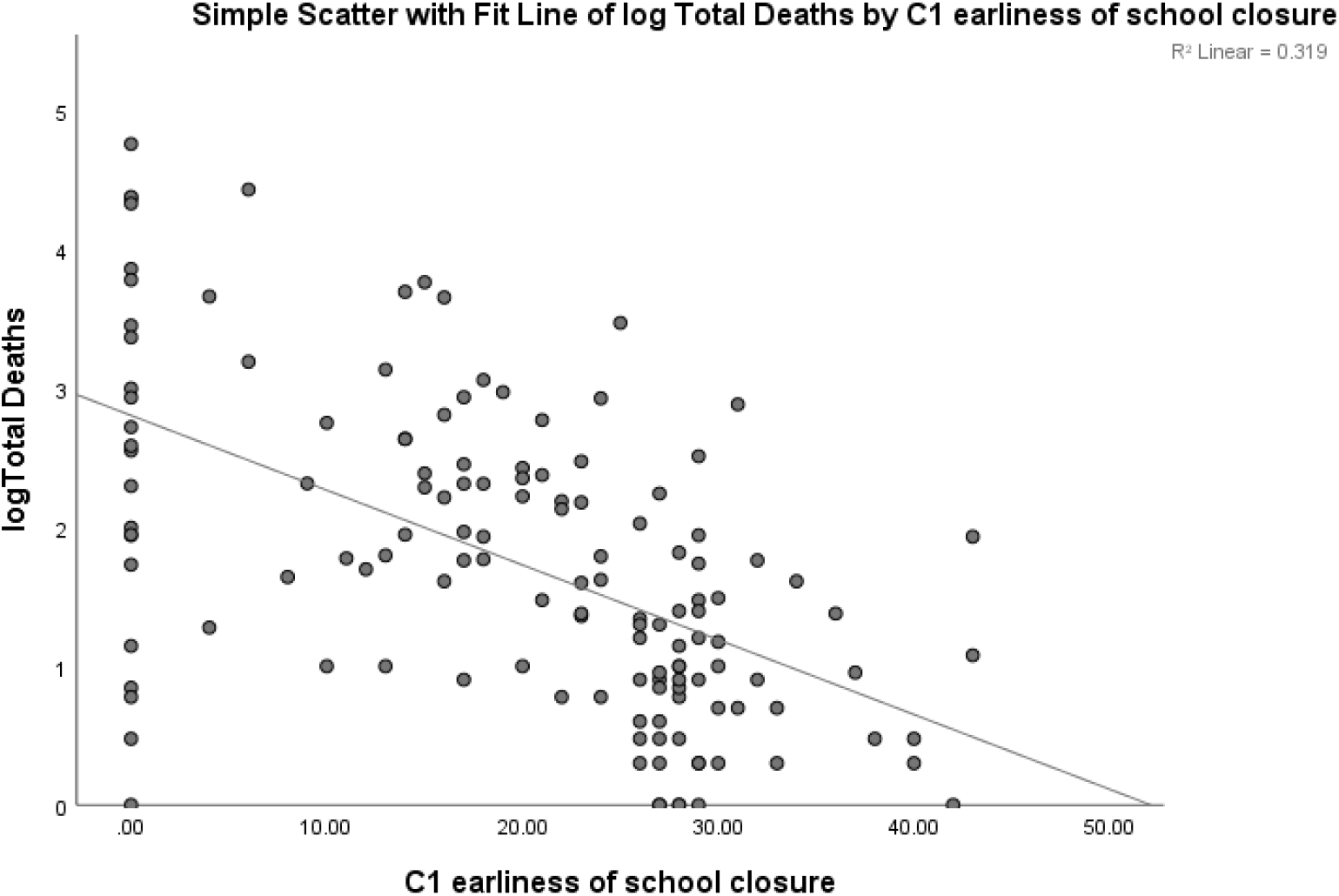
Scatterplot of Log_10_ Total Deaths versus School Closure Earliness as a count back form 28 days after first case.

**Figure 2:**
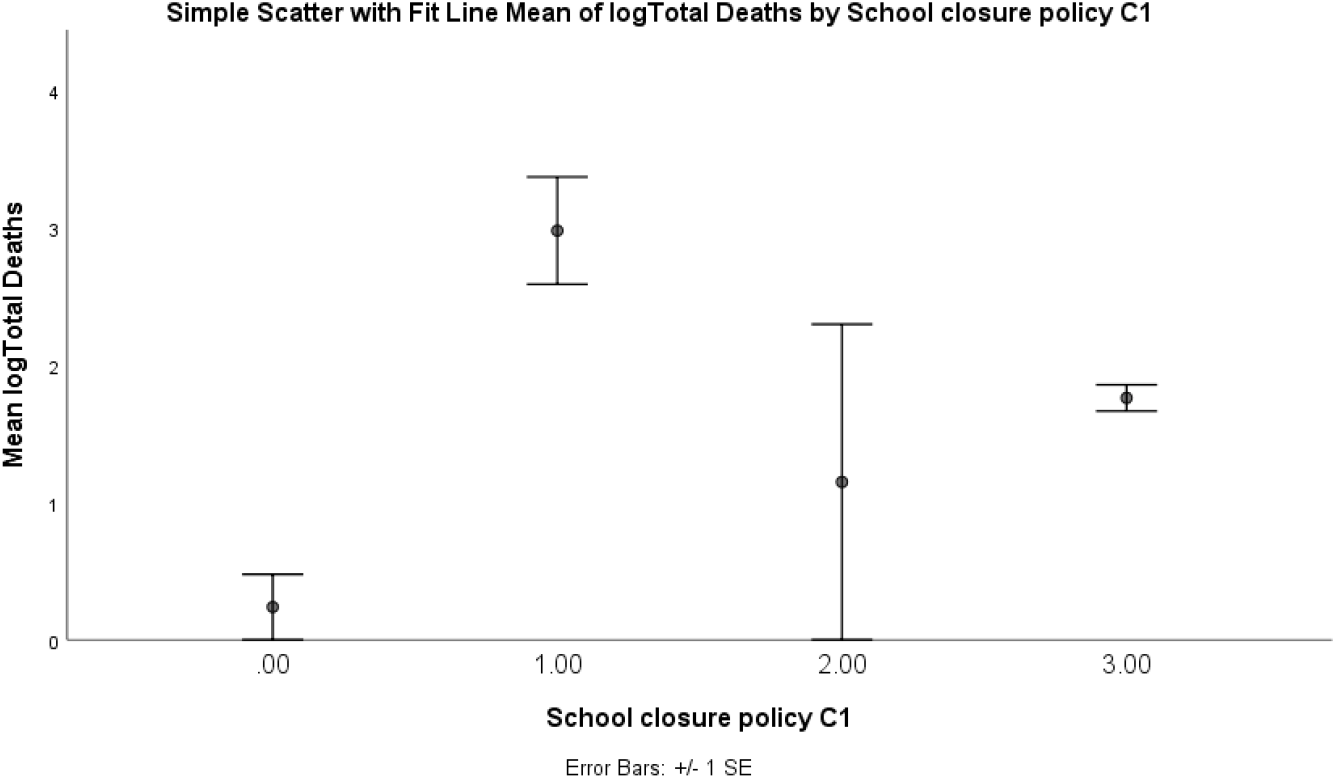
Log_10_ Total Deaths versus Maximum stringency of School Closure Policy. 0: no measures, 1: recommend closing, 2: Require closing (only partially) 3: Require closing (all levels).

A multivariate model was created using demographic factors. Using backwards elimination of factors, the final model included Log of population, purchasing power, life expectancy, date of first COVID-19 death, longitude, average household size, and had an r^2^=0·72 for LogD and r^2^=0·74 for LogC (Table 4). These variables were used in a separate assessment of the timing of introduction of each lockdown policy (Table 5). Early public information (p=0·021, β=-0·005 (95% CI -0·001, -0·009)), early first lockdown measure (p=0·048, β=-0·004 (95% CI 0,-0·008)) and early international travel restrictions (p=0·042, β=-0·005 (95%CI -0·001, -0·009)) were significantly associated with reduced case numbers (LogC). There was a trend for early workplace closure (p=0·094, β=-0·009 (95% CI 0·001, -0·019)) and early generalisation of workplace closure (p=0·052, β=-0·008 (95% CI 0, -0·016)) being associated with LogC. Early public information (p= 0·003, β=-0·007 (95% CI -0·003,-0·011)), and generalised public information policy (p=0·034, β=-0·005 (95%CI -0·001,-0·009)), early international travel restrictions (p=0·003, β=-0·008 (95%CI -0·004, -0·012)), early introduction of first lockdown measure (p=0·003, β=-0·007 (95%CI -0·003,-0·011)), early generalisation of workplace closure (p=0·031, β=-0·012 (95%CI =-0·002, -0·022)) and early generalisation of school closure (p=0·050, β=- 0·012 (95% CI 0, -0·024)) were associated with reduced mortality. There was a trend for early workplace closure (p=0·095, β=-0·011 (95% CI 0·001,-0·023)) being associated with reduced mortality. Throughout Table 5 the coefficients for regression are almost universally negative indicating a prevailing trend for early introduction of policies being beneficial.

**Table 4.** Summary of final demographic variables that were subsequently used in mutlivariate regression to evaluate the correlation between policy timing and Log_10_ Total cases and Log_10_ Total Deaths. The evaluation here is for predcition of log_10_ total deaths (r^2^ =0·72).

|  | Unstandardized Coefficients |  | Standardized Coefficients |  | t | Sig. |
| --- | --- | --- | --- | --- | --- | --- |
|  | B | Std. Error | Beta |  |  |  |
| (Constant) | -5.230 | 1.608 |  |  | -3.253 | 0.001 |
| Log Population | 0.766 | 0.084 | 0.548 |  | 9.065 | 0.000 |
| Purchasing power | 0.034 | 0.027 | 0.072 |  | 1.276 | 0.205 |
| Life expectancy | 0.050 | 0.012 | 0.329 |  | 4.071 | 0.000 |
| Longitude | -0.004 | 0.001 | -0.203 |  | -3.922 | 0.000 |
| 1<sup>st</sup> death | -0.020 | 0.005 | -0.266 |  | -3.912 | 0.000 |
| Household size | -0.144 | 0.056 | -0.173 |  | -2.583 | 0.011 |

**Table 5:** Multivariate regression of timing of policy introduction versus Log_10_ Total Cases and Log_10_ Total Deaths. *= P≤0.05, **= P ≤0.01.

|  | Coefficient Beta<br>Log Total Cases<br>(95% CI) | Sig. (2-tailed)<br>n=133 | Coefficient Beta<br>Log Total Deaths<br>(95% CI) | Sig. (2-tailed),<br>n=122 |
| --- | --- | --- | --- | --- |
| C1 school closing timing | -0.007 | ns | -0.008 | ns |
| C1 timing of general policy | -0.006 | ns | -0.012<br>(0, -0.024) | 0.050 |
| C2 workplace closing timing | -0.009<br>(0.001, -0.019) | 0.094 | -0.011<br>(0.001, -0.023) | 0.095 |
| C2 general policy timing | -0.008<br>(0, 0.016) | 0.052 | -0.012<br>(-0.002, -0.022) | 0.031 |
| C3 cancel public events timing | -0.005 | ns | -0.008 | ns |
| C3 timing of general policy | -0.005 | ns | -0.007 | ns |
| C4 restriction on gatherings timing | 0.001 | ns | 0.000 | ns |
| C4 timing of general policy | -0.003 | ns | -0.001 | ns |
| C5 close public transport timing | 0.006 | ns | 0.004 | ns |
| C5 timing of general policy | 0.000 | ns | 0.003 | ns |
| C6 stay at home restriction timing | -0.004 | ns | -0.003 | ns |
| C6 general policy timing | -0.005 | ns | -0.003 | ns |
| C7 domestic travel restriction timing | -0.004 | ns | -0.007 | ns |
| C7 general policy timing | -0.009<br>(0.001, -0.019) | 0.066 | -0.008 | Ns |
| C8 international travel restriction timing | -0.005<br>(-0.001, -0.009) | 0.042 | -0.008<br>(-0.004, -0.012) | 0.003 |
| H1 public information timing | -0.005<br>(-0.001, -0.009) | 0.021 | -0.007<br>(-0.003, -0.011) | 0.003 |
| H1 timing of general policy | -0.003 | ns | -0.005<br>(-0.001, -0.009) | 0.034 |
| H2 testing policy timing | 0.001 | ns | -0.002 | ns |
| H3 contact tracing timing | 0.002 | ns | -0.001 | ns |
| First lockdown policy introduction | -0.004<br>(0, -0.008) | 0.048 | -0.007<br>(-0.003, -0.011) | 0.003 |

## Discussion

We have presented the first study examining the effect of individual lockdown policies at a national level and found that earlier implementation of some policies; school closure, workplace closure, public information and restriction on foreign travel, were associated with reduced COVID-19 mortality. It was considered beneficial to analyse this data, before the pandemic has taken its course and mortality can be counted more accurately as many decisions about relaxing the lockdown are being considered at this time that may have lasting consequences.^19^

Making comparison between nations may be compromised by variation in testing policies, case reporting, definitions, uptake of policies and medical care, and there is no accounting for these factors here as the real population incidence and the accuracy with which it has been assessed is unknown in any country. Nonetheless we believe a comparison should be undertaken, as actual data should be used to test policies that have been drawn up by conceptual models. Despite these limitations, the model used here has a high concordance with observed data with r^2^=0·72 for LogD and 0·74 for LogC. More precise models with r^2^=0·84 could have been achieved with further factors, at the expense of loss of data and overloading competing COVID-19 responses which may be associated with each other. As much as possible in the multivariate model, we aimed to only include pre pandemic demographics as predictive factors, though it was necessary to include the date of the first death within a country. One of the difficulties with the comparison is that, at this time, both the intervention and the outcome are time dependent. We sought to account for the timing of intervention by counting it within the same time frame for every country and for the timing of the outcome by using the date of the first death as a factor, although it may have been determined in part by the COVID-19 policy. The model is only of utility at one point in time, is in no way predictive and only served to test the lockdown policies.

The size of the effect here from the multivariate regression, taking closing of schools as an example, is such that implementing the policy 24 days earlier was associated with halving of the mortality as of the 29^th^ of April 2020. The measurable effect of introducing several policies together would be less than additive, as their introduction is associated with each other, though if the 3 most statistically significant factors (generalisation of workplace closure, international travel restrictions and public information) are combined on multivariate regression, the additive effect coefficient B is - 0·026 per day on a Log_10_ scale, equivalent to halving mortality for every 11·6 days earlier that the combined policy was introduced. Comparing this to a theoretical model In simple terms (excluding migration and acquired immunity); taking pre lockdown viral reproductive rate (R_0_) as 2·6 and the serial interval as 5 days from estimates,^20^ introducing a strategy that will reduce R^0^ to 0·8, 11·6 days earlier, would decrease new transmissions over that period by 4·7 times. In theory, for every week out of lock down at R_0_ at 2·6, it would take 4 weeks of lock down at R_0_ =0·8 to revert the viral transmission rate. In fact, after 7 days of pre lockdown at R_0_ =2·6, even reducing and holding R_0_ at 0 perpetually, always result in higher total case number than starting with a perpetual lockdown at R_0_=0·8 at day 1. In theory starting a lockdown earlier (provided R_0_<1), has a much greater effect on reducing case numbers than the stringency of the lockdown, explaining the inability of this study to detect any correlation between stringency and outcome at this point in time. It is also possible that starting a lockdown earlier could facilitate a shorter lockdown and thereby provide economic benefits in addition to reducing mortality. Off course perpetual lock downs are theoretical and politically non-viable and the gains from any effective lockdown could be undone by the exit strategy, and in the absence of population immunity, prove to be only temporary. The exit strategy is dependent on the aims of the lockdown which could range from eradicating the disease, to delaying the disease to when a vaccine or suitable therapies become available, to reducing serious case prevalence to below the national ventilator capacity. This final aim does not reduce the ultimate number of cases. The long-term persistence of COVID-19 is unknown, but it is anticipated that there will be a resurgence or second peak which is expected to be less fatal than the first.^19^ The H1N1 1928 “Spanish Flu” outbreak caused 50 million deaths worldwide, in three extensive pandemic waves with the second one being particularly more dramatic.^21^ H1N1 was also considered “novel”, as no person had any immunity in that era. Transmission was similar to Sar-Cov-2 and no vaccines or effective treatment were available. H1N1 showed higher mortality rates to healthy individuals between 20 and 40 years old, compared to SARS-CoV-19 that tends to affect people above 65 or with several comorbidities.

The lockdown measures that we have found to be most effective; school and workplace closure and international travel restrictions have huge political and economic ramifications. Already the effects on transports systems, education, agricultural and manufacturing industries have been measured in colossal financial and workforce numbers. Primary sector demonstrated a 20% drop in agricultural commodities.^22,23^ Oil demand has diminished; by the end of March, barrel prices faced the sharpest drop described over the last 30 years of more than 30%.^24^ Secondary and tertiary sectors have been affected with a reduction of manufacturing companies. Several countries have converted regular teaching to virtual network classes, immediately resulting in 27% of parents being unable to work and an 18% income loss. Numerous airlines, shipping and transport companies have provisionally cancelled their services.^25^ Although the pathogen has not completed its tour, it is estimated to cost as much as 4·1 trillion USD which reflects 5% of the global gross domestic product. The above figure could be much greater if the financial impact on tourism, consumption and investment are accounted for.^26,27^ An estimated 1·6 billion workers, almost half of the global workforce, may face financial difficulties in the second half of 2020. The pre-crisis 6·7% working-hours loss estimate has now escalated to 10·5% reduction due to prolongation of lockdown measures.^28^ Liquidity support for sectors facing disruptions accounts for 16% of EU GDP with 3% of EU GDP in fiscal measures.^29^ Likewise, 540 billion euros support was directed to businesses amongst member states, aiming to minimize unemployment and enable workers to keep their jobs.^30^

Sustaining the lockdown to a point in time when a vaccine may be available is considered by many nations to be untenable, and the conundrum of how to withdraw or minimise lockdown policies is being widely considered.

Given enough time, a lock down that results in R_0_<1, should in theory lead to a state of negligible transmission. Several countries that have entered the pandemic at an early stage, have reported measured community transmission close to zero, which has persisted even after several weeks of lockdown easing combined with tight international travel control^1^, indicating a benchmark which may be replicated. What this means in the fullness of time is still to be determined.

## Data Availability

Data Sharing
We collected data from publically available databases. Our collated study database will be made freely available from the date of journal publication, upon request at

## Conclusions

At this stage in the pandemic, early institution of public information, international travel restrictions, and workplace closure are associated with reduced COVID-19 mortality and maintaining these policies may help control the pandemic.

## Contributors

SB and DIP wrote the manuscript and conducted literature searches

SB designed the study, collected data. Analysed the data and interpreted the result

ID and KC wrote the manuscript

## Data Sharing

We collected data from publically available databases. Our collated study database will be made freely available from the date of publication, upon request at

## References

1. Johns Hopkins Coronavirus Resource Center [Internet]. [cited 2020 May 3]. Available from: https://coronavirus.jhu.edu/

2. The economic impact of COVID-19 | Research | University of Oxford [Internet]. [cited 2020 May 3]. Available from: https://www.research.ox.ac.uk/Article/2020-04-07-the-economic-impact-of-covid-19

3. Yasmin A. MALIK. Properties of Coronavirus and SARS-CoV-2. Malaysian J Pathol. 2020 Apr 11;42(1):3–11.

4. van Doremalen N, Bushmaker T, Morris DH, Holbrook MG, Gamble A, Williamson BN, et al. Aerosol and Surface Stability of SARS-CoV-2 as Compared with SARS-CoV-1 [Internet]. Vol. 382, The New England journal of medicine. NLM (Medline); 2020 [cited 2020 May 3]. p. 1564–7. Available from: http://www.nejm.org/doi/10.1056/NEJMc2004973

5. Huang C, Wang Y, Li X, Ren L, Zhao J, Hu Y, et al. Clinical features of patients infected with 2019 novel coronavirus in Wuhan, China. The Lancet. 2020 Feb 15;395(10223):497–506.

6. Li Q, Guan X, Wu P, Wang X, Zhou L, Tong Y, et al. Early Transmission Dynamics in Wuhan, China, of Novel Coronavirus–Infected Pneumonia. New England Journal of Medicine [Internet]. 2020 Mar 26 [cited 2020 May 3];382(13):1199–207. Available from: http://www.nejm.org/doi/10.1056/NEJMoa2001316

7. Bi Q, Wu Y, Mei S, Ye C, Zou X, Zhang Z, et al. Epidemiology and transmission of COVID-19 in 391 cases and 1286 of their close contacts in Shenzhen, China: a retrospective cohort study. The Lancet Infectious Diseases [Internet]. 2020 Apr [cited 2020 May 3];0(0). Available from: https://linkinghub.elsevier.com/retrieve/pii/S1473309920302875

8. Tu Y-F, Chien C-S, Yarmishyn AA, Lin Y-Y, Luo Y-H, Lin Y-T, et al. A Review of SARS-CoV-2 and the Ongoing Clinical Trials. International Journal of Molecular Sciences [Internet]. 2020 Apr 10 [cited 2020 May 3];21(7):2657. Available from: https://www.mdpi.com/1422-0067/21/7/2657

9. Cheng VCC, Wong S-C, Chuang VWM, So SYC, Chen JHK, Sridhar S, et al. The role of community-wide wearing of face mask for control of coronavirus disease 2019 (COVID-19) epidemic due to SARS-CoV-2. Journal of Infection. 2020 Apr;

10. Sen-Crowe B, McKenney M, Elkbuli A. Social distancing during the COVID-19 pandemic: Staying home save lives. American Journal of Emergency Medicine. W.B. Saunders; 2020.

11. Giordano G, Blanchini F, Bruno R, Colaneri P, di Filippo A, di Matteo A, et al. Modelling the COVID-19 epidemic and implementation of population-wide interventions in Italy. Nature Medicine. 2020 Apr 22;1–6.

12. di Gennaro F, Pizzol D, Marotta C, Antunes M, Racalbuto V, Veronese N, et al. Coronavirus Diseases (COVID-19) Current Status and Future Perspectives: A Narrative Review. International Journal of Environmental Research and Public Health [Internet]. 2020 Apr 14 [cited 2020 May 3];17(8):2690. Available from: https://www.mdpi.com/1660-4601/17/8/2690

13. Coronavirus Government Response Tracker | Blavatnik School of Government [Internet]. [cited 2020 May 3]. Available from: https://www.bsg.ox.ac.uk/research/research-projects/coronavirus-government-response-tracker

14. Household: Size and Composition 2018 - Countries [Internet]. [cited 2020 May 3]. Available from: https://population.un.org/Household/index.html#/countries/840

15. Courtemanche C, Garuccio J, Le A, Pinkston J, Yelowitz A. Strong Social Distancing Measures In The United States Reduced The COVID-19 Growth Rate. Health Affairs [Internet]. 2020 May 14 [cited 2020 May 15];10·1377/hlthaff. Available from: http://www.healthaffairs.org/doi/10.1377/hlthaff.2020.00608

16. Manchein C, Brugnago EL, da Silva RM, Mendes CFO, Beims MW. Strong correlations between power-law growth of COVID-19 in four continents and the inefficiency of soft quarantine strategies. Chaos: An Interdisciplinary Journal of Nonlinear Science [Internet]. 2020 Mar 31 [cited 2020 May 15];30(4):041102. Available from: http://arxiv.org/abs/2004·00044

17. Munayco C v., Tariq A, Rothenberg R, Soto-Cabezas GG, Reyes MF, Valle A, et al. Early transmission dynamics of COVID-19 in a southern hemisphere setting: Lima-Peru: February 29th–March 30th, 2020. Infectious Disease Modelling [Internet]. 2020 May 12 [cited 2020 May 15]; Available from: https://linkinghub.elsevier.com/retrieve/pii/S2468042720300130

18. Colbourn T. COVID-19: extending or relaxing distancing control measures. Vol. 0, The Lancet Public Health. Elsevier Ltd; 2020.

19. Xu S, Li Y. Beware of the second wave of COVID-19. Vol. 395, The Lancet. Lancet Publishing Group; 2020. p. 1321–2.

20. Park M, Cook AR, Lim JT, Sun Y, Dickens BL. A Systematic Review of COVID-19 Epidemiology Based on Current Evidence. Journal of Clinical Medicine [Internet]. 2020 Mar 31 [cited 2020 May 14];9(4):967. Available from: https://www.mdpi.com/2077-0383/9/4/967

21. Taubenberger JK, Morens DM. 1918 Influenza: The mother of all pandemics. Vol. 12, Emerging Infectious Diseases. Centers for Disease Control and Prevention (CDC); 2006. p. 15–22.

22. Prices of agricultural commodities drop 20% post COVID-19 outbreak - Rediff Realtime News [Internet]. [cited 2020 May 3]. Available from: https://realtime.rediff.com/news/india/Prices-of-agricultural-commodities-drop-20-post-COVID19-outbreak/955078599584b749?src=interim_also

23. Xconomy: Fears Of Recession Mount As Coronavirus Declared A Pandemic [Internet]. [cited 2020 May 3]. Available from: https://xconomy.com/national/2020/03/19/fears-of-recession-mount-as-coronavirus-declared-a-pandemic/

24. Oil Prices, Stocks Plunge After Saudi Arabia Stuns World With Massive Discounts : NPR [Internet]. [cited 2020 May 3]. Available from: https://www.npr.org/2020/03/08/813439501/saudi-arabia-stuns-world-with-massive-discount-in-oil-sold-to-asia-europe-and-u-?t=1588362702942&t=1588517540655

25. Coronavirus response | Mobility and Transport [Internet]. [cited 2020 May 3]. Available from: https://ec.europa.eu/transport/coronavirus-response_en

26. Coronavirus Pandemic Could Cost the World $4·1 Trillion | Time [Internet]. [cited 2020 May 3]. Available from: https://time.com/5814933/coronavirus-pandemic-cost-4-trillion/

27. The Price of the Coronavirus Pandemic | The New Yorker [Internet]. [cited 2020 May 3]. Available from: https://www.newyorker.com/magazine/2020/04/20/the-price-of-the-coronavirus-pandemic

28. COVID-19 Pandemic in the World of Work: ILO Monitor: COVID-19 and the world of work. 3rd Edition [Internet]. [cited 2020 May 3]. Available from: https://www.ilo.org/global/topics/coronavirus/impacts-and-responses/WCMS_743146/lang--en/index.htm

29. The EU’s financial response to coronavirus is the biggest in the world | World Economic Forum [Internet]. [cited 2020 May 3]. Available from: https://www.weforum.org/agenda/2020/04/european-union-finance-fiscal-money-support-covid-coronavirus/

30. Report on the comprehensive economic policy response to the COVID-19 pandemic - Consilium [Internet]. [cited 2020 May 3]. Available from: https://www.consilium.europa.eu/en/press/press-releases/2020/04/09/report-on-the-comprehensive-economic-policy-response-to-the-covid-19-pandemic/

